# Potential gains in health-adjusted life expectancy by reducing burden of non-communicable diseases: a population-based study

**DOI:** 10.1101/2022.04.04.22273392

**Authors:** Jun-Yan Xi, Wang-Jian Zhang, Zhuo Chen, Yan-Ting Zhang, Li-Chang Chen, Yu-Qin Zhang, Xiao Lin, Yuan-Tao Hao

## Abstract

**Background:** The United Nations Sustainable Development Goals (SDGs) target 3.4 aims to reduce premature mortality attributable to non-communicable diseases (NCDs) by one-third of their 2015 levels by 2030. Although meeting this target leads to longevity, survivors may suffer from long-term disability caused by NCDs. This paper quantifies the potential gains in health-adjusted life expectancy for people aged 30-70 years (HALE_[30–70)_) by examining the reductions in disability in addition to premature mortality. Additionally, we also assessed the feasibility of meeting the SDGs target 3.4.

**Methods:** We extracted data from the Global Burden of Disease Study 2019 for all NCDs and four major NCDs (cancers, cardiovascular diseases, chronic respiratory diseases, and diabetes mellitus) in 188 countries from 1990 to 2019. Bayesian age-period-cohort models were used to predict possible premature mortality in 2030. The life table was used to estimate the unconditional probability of death and HALE_[30–70)_. Estimates of the potential gains in HALE_[30–70)_ were based on three alternative future scenarios: a) eliminating all premature deaths and disability from a specific cause, b) meeting SDGs target 3.4, and survivors’ disability is eliminated, and c) meeting SDGs target 3.4, but survivors remain disabled for the rest of their lives.

**Results:** In 2030, the unconditional probability of premature mortality for four major NCDs in most countries remained at more than two-thirds of the 2015 baseline. In all scenarios, the high-income group has the greatest potential gains in HALE_[30–70)_, above the global average of HALE_[30–70)_. In scenario A, the potential gains in HALE_[30–70)_ of reducing premature mortality for four major NCDs are significantly lower than those for all NCDs (range of difference for all income groups: 2.88 - 3.27 years). In scenarios B and C, the potential gains of HALE_[30–70)_ in reducing premature mortality for all NCDs and the four major NCDs are similar (scenario B: 0.14 - 0.22, scenario C: 0.05 - 0.19). In scenarios A and B, countries from the high-income group have the greatest potential gains in HALE_[30–70)_ from cancer intervention, whilst countries from the other income groups result in a greater possible HALE_[30–70)_ gains from cardiovascular diseases control. In scenario C, countries from each income group have the largest potential gains in HALE_[30–70)_ from diabetes reduction and chronic respiratory diseases prevention.

**Conclusions:** Achieving SDGs target 3.4 remains challenging for most countries. The elimination of disability among the population who benefit from the target could lead to a sizable improvement in HALE_[30–70)_. Reducing premature death and disability at once and attaching equal importance to each to in line with the WHO goal of “leaving no one behind”.

## Introduction

Reducing the burden of disease caused by non-communicable diseases (NCDs) is a critical target of global public health efforts. According to the World Health Statistics 2021 released by the World Health Organization (WHO), seven of the top ten causes of death in 2019 were NCDs, and the proportion of mortalities attributable to NCDs increased from 60.8% in 2000 to 73.6% in 2019^[1]^. WHO focuses on four categories of NCDs, including cardiovascular diseases (CVD), cancers, chronic respiratory diseases (CRD), and diabetes mellitus, which are the leading causes of premature death from NCDs^[2]^. Although the total mortality rate for the four NCDs is declining, the total number of deaths associated with them is still increasing^[1]^. In addition, the impact of NCDs is particularly severe in low- and middle-income countries, where more than three-quarters of global NCD deaths occur. The burden of NCDs in these countries is predicted to continue increasing in the coming decades^[2]^. Reducing premature mortality (the unconditional probability of dying from the four major NCDs between the ages of 30 and 70) by a third from 2015 levels by 2030 is on the United Nations (UN) agenda as Sustainable Development Goals (SDGs) target 3.4^[3]^. The NCD Countdown 2030 Report proposed more comprehensive indicators and committed to leaving no one behind^[4,5]^. Although members of the UN General Assembly have stepped up their investment in health and begun to develop framework legislation to strengthen national governance of NCDs, slow progress in NCDs prevention and control may make it difficult to meet SDGs target 3.4^[6-8]^. Trend analysis of regional and country-level premature mortality attributable to NCDs from a global health perspective provides an intuitive assessment of progress towards meeting SDGs target 3.4^[9,10]^. Meeting the ambitious SDGs target 3.4 would have substantial effects on life expectancy globally^[9]^. However, neglected disability attributed to the four major NCDs may lead to unhealthy longevity^[11,12]^. Since 1990, there has been a marked shift towards a greater proportion of burden due to years lived with disability (YLDs) from NCDs^[13]^. Many excluded disabilities are amenable to prevention and treatment, and some NCDs deaths can be postponed. Therefore, the global agenda for promoting human prosperity should be expanded to rehabilitate the disability of survivors who benefit from SDGs target 3.4. Health-adjusted life expectancy can effectively estimate the healthy life span lost due to disability by decomposing life expectancy into healthy and disability components^[14]^. Assessing the potential of reducing disability in survivors to extend health-adjusted life expectancy under the framework of SDGs target 3.4 is key to highlighting the public health and socioeconomic significance of this expanded target.

In this study, we grouped 188 countries according to the World Bank income classifications. We fitted Bayesian age-period-cohort (BAPC) models based on historical premature mortality trends from 1990 to 2019 and assessed the feasibility of meeting SDGs target 3.4 by 2030 based on model projections. In addition, we considered three alternative future scenarios in 2030 to quantify the potential gains in health-adjusted life expectancy between ages 30 and 70 years (HALE_[30–70)_) by reducing disability and premature mortality from non-communicable diseases.

## Methods

### Data sources

We obtained standard epidemiological measures from the Global Burden of Disease Study 2019 (GBD 2019) result for five cause clusters by country and year, including all-cause mortalities or cause-specific mortalities (non-communicable diseases, neoplasms, cardiovascular diseases, chronic respiratory diseases, and diabetes mellitus), demographics, years of life lost (YLLs) and years lived with disability (YLDs). GBD 2019 is a multinational collaborative study that estimates the burden of diseases in countries around the world and produces summary measures of health. The study updates the datasets annually, thus consistent comparisons could be made between sex, age, and across locations for 1990-2019. The data sources and methods are described elsewhere^[13-15]^. All datasets are available via the GBD 2019 website^[16]^.

Population projections for 2020-2030 were obtained from the World Population Prospects 2019. This dataset provides the median, 80%, and 95% prediction intervals with quinquennial population by country, sex, and five-year age groups (0-4, 5-9, 10-14, …, 95-99, 100+). World Population Prospects 2019 is probabilistic population projections produced and displayed by the United Nations Population Division based on the Bayesian hierarchical models for the total fertility rate and life expectancy at birth, with detailed methods reported previously^[17,18]^. The dataset is available online^[19]^.

In addition, we grouped selected countries based on the World Bank’s classification, including high-income countries, upper-middle-income countries, lower-middle-income countries, and low-income countries^[20]^. This study complies with the Guidelines for Accurate and Transparent Health Estimates Reporting (GATHER) statement^[21]^.

### Analytical strategies

The Bayesian APC models were fitted by NCDs cluster, gender, and country to predict possible NCDs-attributable premature mortality in 2030, based on historical trends in premature mortality from 1990 to 2019. The Bayesian APC models, which incorporates age, period, and cohort heterogeneity, were developed with R software, using the BAPC package (R version 4.1.1)^[22]^. BAPC package builds upon Integrated Nested Laplace Approximations (INLA) and addresses complex convergence concerns introduced by Markov chain Monte Carlo (MCMC)^[23]^. The aim is to efficiently forecast future rates and counts within a fully Bayesian inference setting. Specifically, we chose to use second-order random walk priors (RW2) for age, period, and cohort effect, which penalizes deviations from a linear trend with better predictions. The hyper prior for the precision (inverse variance) in the random walk priors is a Gamma distribution with parameters *a* and *b*, and weak hyperparameters (*a*=1, *b*=0.00005) were set to smooth the estimation^[24]^. The dataset used for age-standardization was obtained from GBD 2019 global population estimates.

We estimated the potential gains in HALE_[30–70)_ by applying cause-deleted life table methods to cause-specific baseline premature mortality reported in 2015, using methods developed by Sullivan^[14,25,26]^. The key to estimating the potential gain in HALE_[30–70)_ from a specific cause is to calculate the cause-deleted death probability and cause-deleted disability prevalence, as described in previous studies^[27,28]^, which is calculated assuming independence among causes of death and disability. We considered three alternative future scenarios of premature mortality reduction from what was observed in the baseline year 2015 as in the following.

a. Eliminating all premature deaths and disability from a specific cause to assess the maximum potential to increase HALE_[30–70)_, which reflects all health burdens of the specific cause.

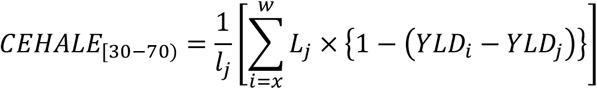
b. Based on the expanded target, the unconditional probability of premature death from a specific cause is reduced by one-third, and survivors’ disability is eliminated, which reflects the potential to increase HALE_[30–70)_.

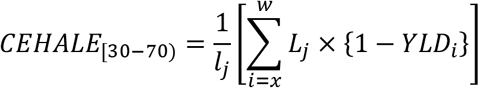
c. Based on SDGs target 3.4, the unconditional probability of premature death from a specific cause is reduced by one-third, but survivors are disabled for the rest of their lives, which reflects the healthy years of life lost to disability due to the specific cause and is used as a reference.

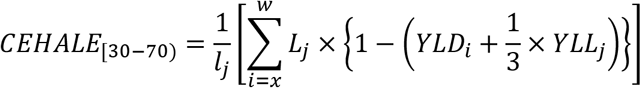

Where *l*_*j*_ is the number of persons surviving to the exact age *x*_*i*_ after eliminating or reducing by a third number of deaths attributed to the cause *j, L*_*j*_ is the number of person-years lived in the age interval (*x*_*i*_, *x*_*i* + 1_) after eliminating the number of person-years attributed to cause *j, YLD*_*i*_ and *YLD*_*j*_ is years lived with disability due to all-cause and specific-cause in the age interval (*x*_*i*_, *x*_*i* + 1_), *YLD*_*j*_ is years of life lost due to specific-cause in the age interval (*x*_*i*_, *x*_*i* + 1_), and *ω* is the last age group in the life table.

The difference between CEHALE_[30–70)_ and HALE_[30–70)_ is the potential gain. We computed the 95% uncertainty intervals (UIs) by propagating the uncertainty from the GBD 2019 estimates.

## Results

### Trends in age-standardized premature mortality for all NCDs and four major NCDs in people aged 30-70, 1990-2030

From 1990 to 2030, the age-standardized premature mortality of all NCDs showed varying degrees of decline (ranging from 24.8% to 45.3%). Specifically, high- and upper-middle-income countries had higher age-standardized premature mortality due to cancers in 1990 than other countries, but the relative decline in this rate over three decades was more than 30 percent (35.1%, 31.9%), higher than lower-middle- and low-income countries (4.5%, 9.9%). The relative decline in age-standardized premature mortality from CVD was more than 50 percent in high- and upper-middle-income countries (58.0%, 53.9%) and close to 30 percent in lower-middle- and low-income countries (27.4%, 29.6%). The relative decline in age-standardized premature mortality attributable to CRD was greatest in upper-middle-income countries (78.6%) and smallest in high-income countries (35.1%), but the latter already had the lowest premature mortality in 1990. Although the global age-standardized premature mortality for the above three major NCDs decreased, the age-standardized premature mortality for diabetes mellitus increased by 9.9%. The age-standardized premature mortality rate for diabetes in other countries showed a downtrend, except for lower-middle-income countries. Age-standardized premature mortality due to cancers is projected to remain higher in high- and upper-middle-income countries until 2030 compared with lower-middle- and low-income countries, while age-standardized premature mortality due to the three major NCDs are expected to be low (figure 1).

**Fig. 1.**
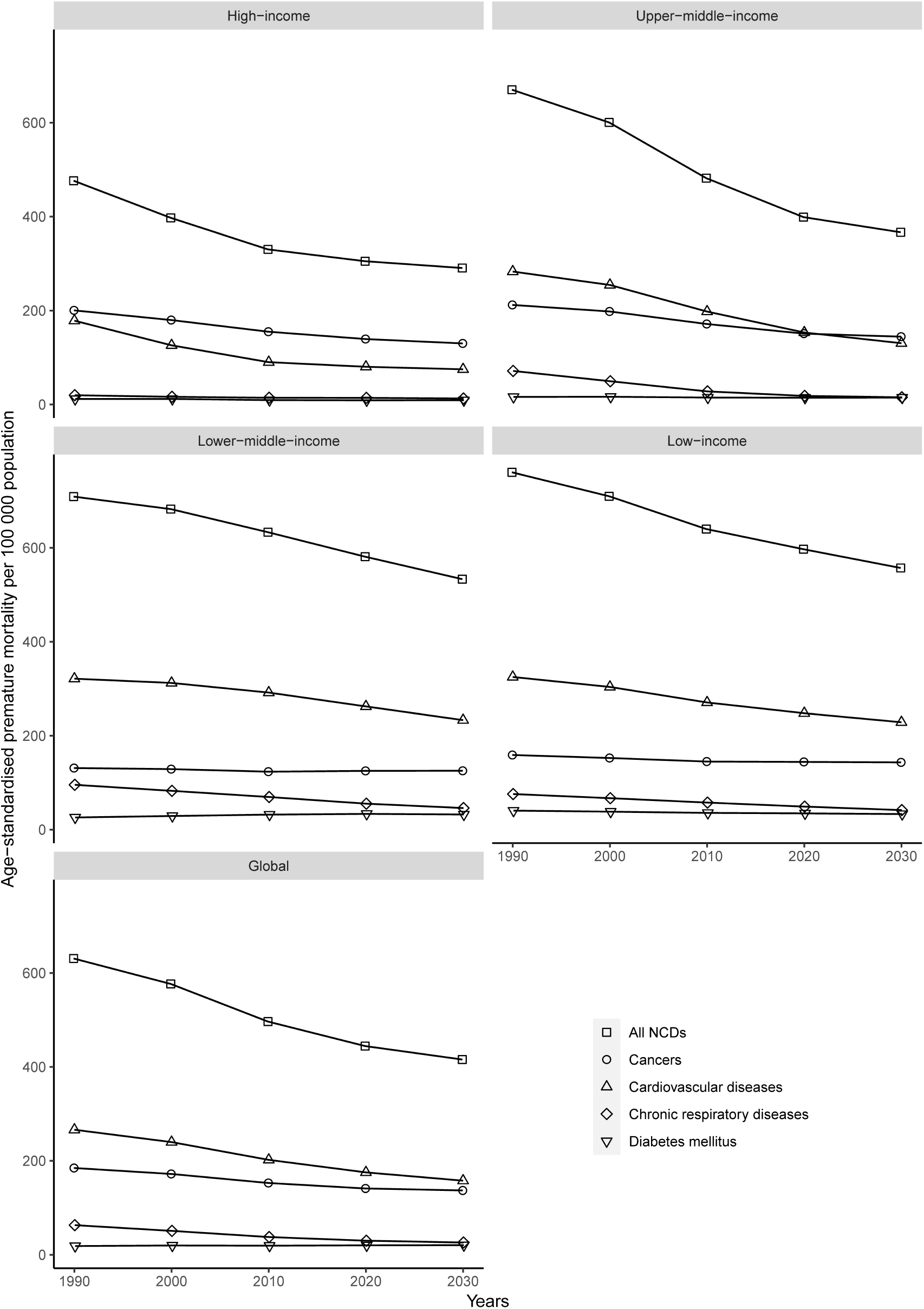
Trends in age-standardized premature mortality for all NCDs and four major NCDs in people aged 30-70, 1990-2030.

### Changes in unconditional probability of premature death for four major NCDs in 2030 compared with 2015

Figure 2 shows the estimated unconditional probability of premature death reduction attributable to four major NCDs by 2030 compared to 2015. The global premature mortality in 2030 is only marginally lower than in 2015 and still far from the goal of reducing it by a third (premature mortality: 0.024 in 2015, 0.023 in 2030; 0.016 in SDGs target 3.4). In 2030, the unconditional probability of premature mortality for four major NCDs in most countries remained at more than two-thirds of the 2015 baseline. While high-income countries will struggle to reduce premature mortality by a third, their baseline for 2015 and projected for 2030 are well below global. For the other three income groups, lower income was associated with higher premature mortality. The unconditional probability of premature death for four major NCDs for 2015 and 2030 and their proportional changes with 95% UIs are provided in the appendix (table S1).

**Fig. 2.**
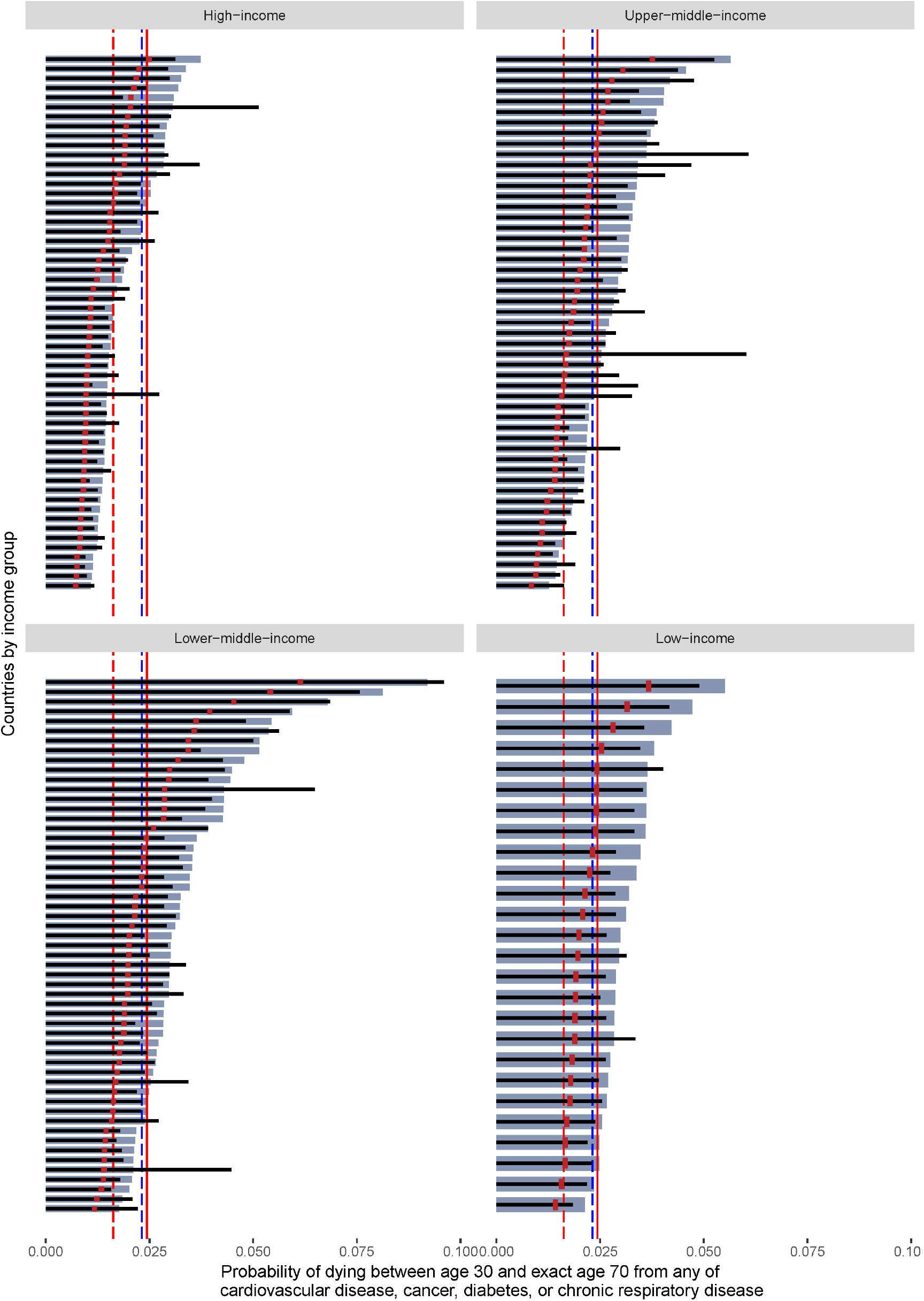
Changes in unconditional probability of premature death for four major NCDs in 2030 compared with 2015.

In the stack bar graph, gray and black columns are the unconditional probabilities of premature mortality for the four major NCDs in 2015 and 2030, respectively. The red dots represent 2/3 of the unconditional probability of premature mortality at baseline in 2015. The red dotted lines, blue dotted lines, and red solid lines represent the SDG target 3.4 reference value, the projected value for 2030, and the baseline value for 2015 respectively.

### HALE_[30–70)_ and age-standardized YLDs for all NCDs and four major NCDs in people aged 30-70, 2015

In 2015, the HALE_[30–70)_ of the global population was 25.14 years of the 40-year lifespan that corresponds to ages 30-70 years. The HALE_[30–70)_ in high- and upper-middle-income countries (28.01 years, 25.71 years) were above the world level, while lower-middle- and low-income countries (22.54 years, 21.35 years) were below it. The proportion of YLDs attributed to all NCDs ranges from 88.2% in high-income countries to 75.1% in low-income countries. Overall, the proportion of YLDs attributed to diabetes mellitus was the highest of the four major NCDs, and the opposite was true for cancers. Cancers had a higher proportion of YLDs in high- and upper-middle-income countries than in lower-middle- and low-income countries. Lower-middle-income countries had the highest proportion of YLDs in CRD. Notably, upper-middle-income countries had the lowest proportion of YLDs for CRD, but the leading cause of YLDs was CVD. On the contrary, the proportion of YLDs in CVD in other countries was lower than the world level (table 1). The average HALE and age-standardized YLDs between 30 years and 70 years with 95% UIs are provided in the appendix (table S2).

**Table 1.** HALE_[30–70)_ and age-standardized YLDs for all NCDs and four major NCDs in people aged 30-70, 2015.

|  | Global | High-income | Upper-middle-income | Lower-middle-income | Low-income |
| --- | --- | --- | --- | --- | --- |
| HALE<sub>[30–70]</sub> | 25.14 | 28.01 | 25.71 | 22.54 | 21.35 |
| Attributable age-standardized YLDs (per 100 000) |  |  |  |  |  |
| All-Cause | 14087.2<br>(100%) | 14646.2<br>(100%) | 12940.2<br>(100%) | 15091.9<br>(100%) | 14839.6<br>(100%) |
| All NCDs | 11808.2<br>(83.8%) | 12916.1<br>(88.2%) | 11169.6<br>(86.3%) | 12113.3<br>(80.3%) | 11140<br>(75.1%) |
| Major NCDs | 1807.8<br>(12.8%) | 1878.0<br>(12.9%) | 1706.1<br>(13.2%) | 1906.2<br>(12.6%) | 1682.6<br>(11.3%) |
| Cancers | 137.7<br>(1.0%) | 244.4<br>(1.7%) | 133.9<br>(1.0%) | 73.9<br>(0.5%) | 68.9<br>(0.5%) |
| CVD | 511.3<br>(3.6%) | 420.2<br>(2.9%) | 563.6<br>(4.4%) | 496.8<br>(3.3%) | 520.6<br>(3.5%) |
| CRD | 463.5<br>(3.3%) | 508.5<br>(3.5%) | 357.4<br>(2.8%) | 576.6<br>(3.8%) | 480<br>(3.2%) |
| Diabetes mellitus | 695.3<br>(4.9%) | 704.9<br>(4.8%) | 651.2<br>(5.0%) | 758.9<br>(5.0%) | 613.1<br>(4.1%) |

HALE<sub>[30–70]</sub> = health adjusts life expectancy for people aged 30-70; YLDs = years lived with disability; NCDs = non-communicable diseases; CVD = cardiovascular diseases; CRD = chronic respiratory diseases

### Potential gains in health-adjusted life expectancy for people aged 30-70 in alternative future scenarios

In all scenarios, the high-income group has the greatest potential gains in HALE_[30–70)_, above the global average. For all specific causes, potential gains in HALE_[30–70)_ decreases as income levels fall. In scenario A, the potential gains in HALE_[30–70)_ of reducing premature mortality for four major NCDs are significantly lower than for all NCDs (range of difference for all income groups: 2.88 - 3.27 years). In scenario B and scenario C, the potential gains of HALE_[30–70)_ in reducing premature mortality for all NCDs and the four major NCDs are very close (scenario B: 0.14 - 0.22, scenario C: 0.05 - 0.19). In scenarios A and B, the high-income group has the greatest potential gains in HALE_[30–70)_ from cancer intervention, and the other income groups have the greatest potential gains in HALE_[30–70)_ from cardiovascular diseases intervention. In scenario C, all income groups have the greatest potential gains in HALE_[30–70)_ from diabetes and chronic respiratory diseases (figure 3).

**Fig. 3.**
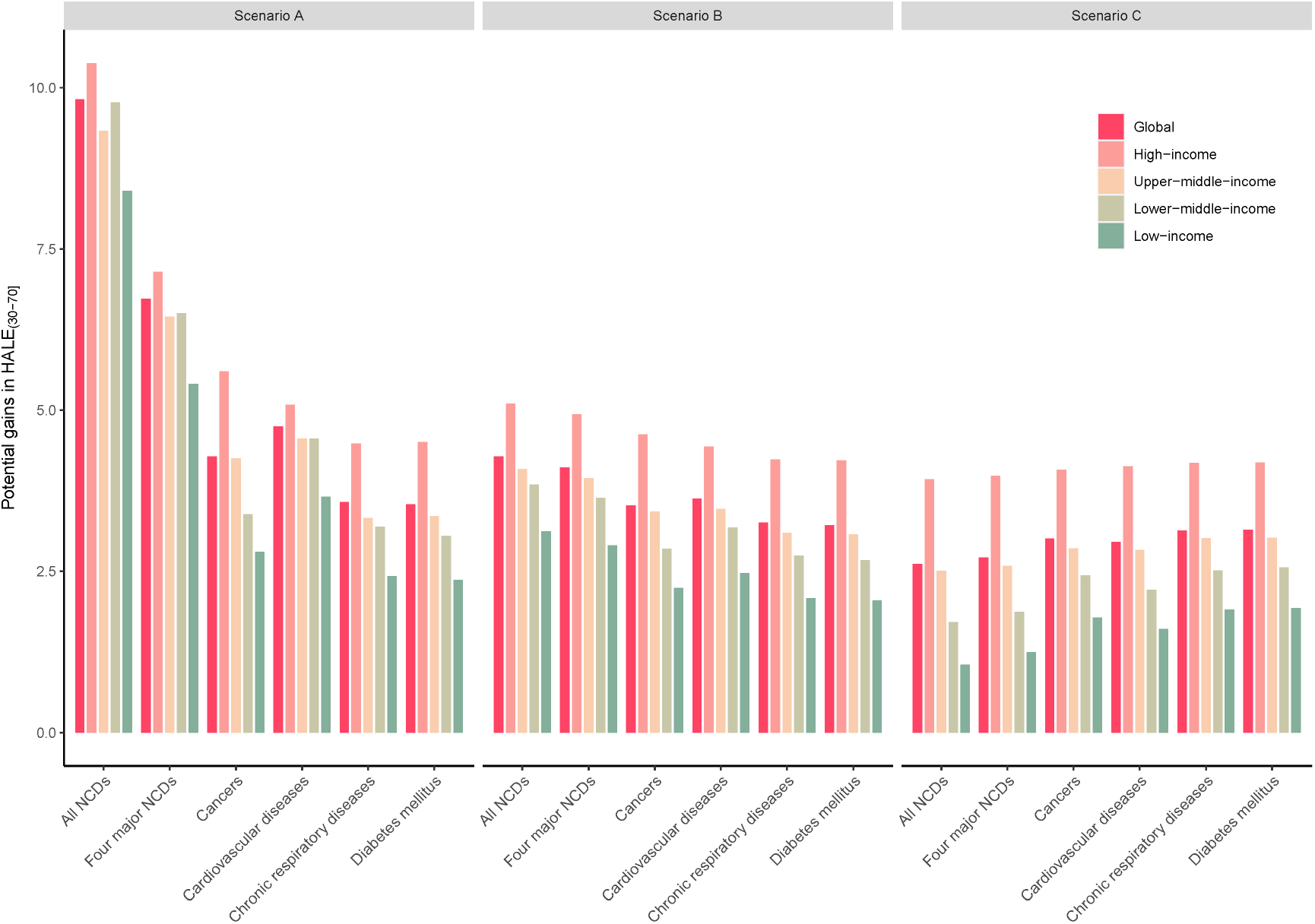
For specific non-communicable diseases, potential gains in health-adjusted life expectancy for people aged 30-70 in alternative future scenarios.

For four major NCDs, reaching scenario A is estimated to yield fewer potential gains of HALE_[30–70)_ in several Southern Africa and Middle Africa regions, including the Central African Republic and Niger. The Pacific island states, including Fiji, Guam, Solomon Islands, and Kiribati, in addition to the United States of America, have the larger benefits. Achieving scenario B is estimated to yield higher potential gains of HALE_[30–70)_ in the United States of America, Australia, Canada, Japan, Iceland, New Zealand, and Kuwait, but some African regions, such as Lesotho and the Central African Republic, have lower potential gains of HALE_[30–70)_. In scenario C, HALE_[30–70)_ will show negative growth in some African and Pacific island states, including Solomon Islands, Kiribati, Uzbekistan, Micronesia (Fed. States of), Lesotho, and the Central African Republic. By contrast, HALE_[30–70)_ is still growing aggressively in certain American, European, and Asian countries, such as Australia, Japan, Iceland, United States of America, Canada, Switzerland, Luxembourg, Republic of Korea, New Zealand, Kuwait, Italy, Norway, Singapore, Colombia, Puerto Rico, Spain, and France (figure 4). For all NCDs, four major NCDs, and the individual major NCDs, potential gains in HALE_[30–70)_ with 95% UIs are provided in the appendix (table S3, table S4, and table S5).

**Fig. 4.**
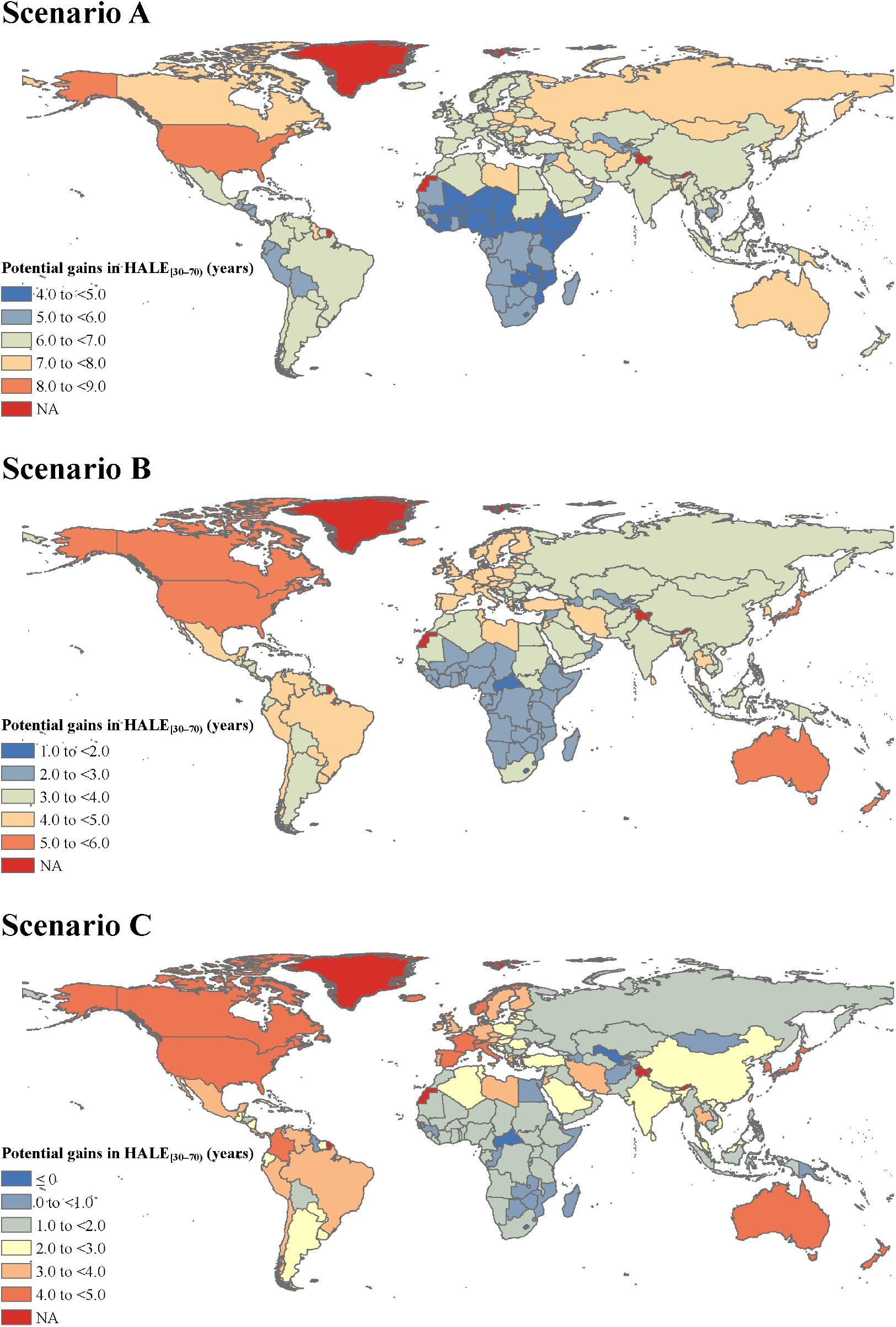
For four major non-communicable diseases, potential gains in health-adjusted life expectancy for people aged 30-70 in alternative future scenarios.

## Discussion

Our results highlight the significant gains in HALE_[30–70)_ worldwide on top of achieving SDGs target 3.4, enabling survivors to rehabilitation from disability, which reflects the current high premature mortality burden and disability burden of NCDs worldwide and the effect that efforts to reduce it could have. However, based on trends in premature mortality over the past three decades, we found that it seems impossible for most countries to reduce premature mortality from four major NCDs to two-thirds of the 2015 baseline within the 10-year window. To our knowledge, this is the first global study to assess the potential gains for HALE_[30–70)_ by reducing premature death and disability as a whole and attaching equal importance to each. This study highlights the limitations of the current framework from a visionary perspective and points toward future directions for necessary global health policy with broadened scope. Our results reinforce the urgency of meeting SGDs target 3.4 and highlight the importance of reducing disability to improve quality of life.

We found HALE_[30–70)_ in high- and upper-middle-income countries were higher than that in lower-middle- and low-income countries in 2015, but the gaps between high- and low-income countries in age-standardized YLDs were small. Current demographic and health risk changes have resulted in an increase in disability and an earlier onset of disability. The increase in the number of total YLDs is significant in the context of population aging and health risk shifts^[29]^. However, a horizontal comparison of HALE based on life expectancy between countries at different economic levels does not reflect the lack of disability rehabilitation, as developed countries have a higher life expectancy^[30]^. The increase in the number of total YLDs may burden economies and healthcare systems as the systems have not kept up with population growth^[31]^. Reducing premature mortality and disability requires enhancing primary care and scale-up rehabilitation, driven by significant population aging and increased demand for long-term care^[32-34]^. Generally, Integrating rehabilitation into primary health care is limited in lower-middle- and low-income countries, while high- and upper-middle-income countries do better^[35-37]^. However, our results suggest that there is still a need for health policies in developed countries to expand the coverage and willingness of rehabilitation and benefit more people. Although 80% of NCDs occurred in lower-middle- and low-income countries^[7]^, we observed that the potential gains in HALE_[30–70)_ from reducing premature mortality and disability for the four major NCDs are highest in high-income countries. The potential contributing factors may be a decrease in poverty-related risk factors and an increase in behavioral risk factors, such as tobacco consumption, harmful use of alcohol, and unhealthy diet^[9,38-41]^. The four major NCDs have received attention due to their high prevalence and high mortality rates. Excessive premature mortality indicates early age of disease, rapid course of the disease, and poor prognosis, and most of them are accompanied by severe complications. Surviving patients must continue to live with the disease, develop new life routines, and adapt to the changes in health status, which means they have a higher disability weight and more disability life years lost. NCDs are long-term or lifelong, and a person cannot hold a temporary “patient role” and then recover. In addition to socio-economic effects, exposure to products that are harmful to health or unhealthy eating habits can also increase the risk of NCDs^[2,42,43]^. NCDs can be controlled by reducing the risk factors associated with these diseases, and guiding policy and priorities by governments and other stakeholders are essential^[2]^. It is recommended that lower-middle- and low-income countries integrate rehabilitation into primary health care, while high- and upper-middle-income countries focus on improving behavioral risk factors while ensuring that sound rehabilitation and care benefit everyone.

Our results revealed estimates of what individual countries in four income groups might achieve in controlling premature mortality in four major NCDs by 2030, which will provide policymakers with experience from leading countries and evidence on health policy. Some European countries are expected to do better in controlling NCDs’ premature mortality, and these countries are considered similar in terms of social structure and culture^[44]^. The Nordic welfare states have a long-standing commitment to egalitarian welfare policies that have helped to achieve significant improvements in the health of covered all social groups^[44,45]^. The WHO European Region has mapped the lessons to the Consolidated Framework for Implementation Research to support increasing coverage of quality essential health services in member states with the lowest resources^[46]^. In lower-middle- and low-income countries, priorities for the prevention and control of NCDs compete with other important agendas such as poverty eradication and environmental protection. It is encouraging that the UN and WHO are actively working to combat the disease burden of NCDs in less developed regions and that NCDs-related strategies such as Universal Health Coverage and Best Buy are integrated into primary health care^[47]^. But few countries met the recommended levels, and there is a lack of adequate evaluation of many well-developed interventions^[48,49]^. NCDs are killing and disabling people at their peak productivity and are already another “poverty trap” on par with infectious diseases^[50]^. A substantial proportion of NCDs among adults could be prevented by early screening and self-management, but lower-middle- and low-income countries appear to have not fully adopted cost-effective strategies, possibly limited by the affordability and accessibility of health care^[37,51]^.

This study has several strengths. First, we provided a trend analysis of regional and country-level NCDs from a global health perspective and an intuitive assessment of progress towards meeting SDGs target 3.4. Secondly, we expanded on SDGs target 3.4 of focusing on reducing disability in survivors and reducing premature mortality, which is a key step toward reducing the disease burden of NCDs. Third, the data sources for our study were estimates modeled from the best available evidence and research, which be viewed as the best current estimate. Open access databases of GBD across locations and the entire time series will be re-estimated by relevant study collaborators each new cycle using the latest datasets and more advanced modeling methods.

This study has at least two limitations. First, for individual NCDs, our analysis did not consider causes beyond the four main NCDs. We limited our analysis to four major NCDs because they are the leading cause of premature death, in line with the SDG target 3.4 emphasizes reducing premature death. Further studies will take into account more NCDs with low mortality but a higher disease burden for survivors. Second, in the hypothetical scenario of estimating potential gains in HALE_[30–70)_, we converted YLLs to YLDs in equal proportions. For some causes and their sequelae, current medical techniques may be difficult to relieve unbearable physical and mental suffering while extending life. Disability weights and YLDs may be underestimated. Therefore, our results are conservative estimates of potential gains of HALE_[30–70)_.

In conclusion, our findings show that meeting SDGs target 3.4 remains challenging for most countries. Currently, the health life-year loss due to NCDs is highest in high-income countries. The elimination of survivor disability from the SDG target 3.4 could lead to a sizable improvement in average health-adjusts life expectancy lived for people aged 30 to70 years by 2030. We call on governments to make robust public health interventions and adequate consideration of the independent or combined effects of health drivers, including health risk factors, interventions, and broader sociodemographic and health system factors, to ensure that health resources are properly distributed and available to all, especially to those most in need^[52]^.

## Data Availability

The full dataset in this study can be obtained from the Institute for Health Metrics and Evaluation (https://www.healthdata.org/) and World Population Prospects 2019 (https://population.un.org/wpp/). All data generated during this study are included in the manuscript and supporting files.

## Acknowledgments

The most important acknowledgment is to the GBD 2019 Diseases and Injuries Collaborators, as well as to all the participants included in the study. The authors alone are responsible for the views expressed in this article and they do not necessarily represent the views, decisions, or policies of the institutions with which they are affiliated.

## Author contributions

Jun-Yan Xi: Conceptualization; Data curation; Software; Formal analysis; Investigation; Visualization; Methodology; Writing - original draft. Wang-Jian Zhang: Formal analysis; Validation; Writing - review and editing. Zhuo Chen: Formal analysis; Validation; Writing - review and editing. Yan-Ting Zhang: Investigation; Visualization; Methodology. Li-Chang Chen: Investigation; Visualization; Methodology. Yu-Qin Zhang: Investigation; Visualization; Methodology. Xiao Lin: Conceptualization; Resources; Supervision; Funding acquisition; Methodology; Project administration; Writing - review and editing. Yuan-Tao Hao: Conceptualization; Resources; Supervision; Funding acquisition; Methodology; Project administration; Writing – review and editing.

## Ethics

The requirement for ethical board approval was waived because this study is secondary data analysis. All data were aggregated and did not contain any information at the individual levels. Therefore, there were no specific ethical issues warranted special attention.

## Funding

**Guangdong Basic and Applied Basic Research Foundation (2020A1515011294)**

Yuan-Tao Hao

**Guangdong Basic and Applied Basic Research Foundation (2020A1515110230)**

Xiao Lin

**Guangdong Basic and Applied Basic Research Foundation (2021A1515011765)**

Xiao Lin

**China Postdoctoral Science Foundation (grant 2021M693594)**

Xiao Lin

Additionally, Professor Yuan-Tao Hao gratefully acknowledges the support of K.C.Wong Education Foundation.

The funders had no role in study design, data collection and interpretation, or the decision to submit the work for publication.

## References

1. World Health Statistics. World Health Statistics 2021. https://apps.who.int/iris/bitstream/handle/10665/342703/9789240027053-eng.pdf. accessed December 1, 2021

2. World Health Organization. Noncommunicable diseases. https://www.who.int/news-room/fact-sheets/detail/noncommunicable-diseases. accessed December 1, 2021

3. United Nations. Transforming our World: The 2030 Agenda for Sustainable Development. https://sustainabledevelopment.un.org/post2015/transformingourworld/publication. accessed December 1, 2021

4. NCD Countdown Collaborators. NCD Countdown 2030. https://ncdcountdown.org/. accessed December 1, 2021

5. Bennett JE, Stevens GA, Mathers CD, et al. NCD Countdown 2030: worldwide trends in non-communicable disease mortality and progress towards Sustainable Development Goal target 3.4. The Lancet. 2018; 392: 1072–88.

6. Nugent R, Bertram MY, Jan S, et al. Investing in non-communicable disease prevention and management to advance the Sustainable Development Goals. The Lancet. 2018; 391: 2029–35.

7. Niessen LW, Mohan D, Akuoku JK, et al. Tackling socioeconomic inequalities and non-communicable diseases in low-income and middle-income countries under the Sustainable Development agenda. The Lancet. 2018; 391: 2036–46.

8. Magnusson RS. Framework legislation for non-communicable diseases: and for the Sustainable Development Goals? BMJ Global Health. 2017; 2: e385.

9. Cao B, Bray F, Ilbawi A, et al. Effect on longevity of one-third reduction in premature mortality from non-communicable diseases by 2030: a global analysis of the Sustainable Development Goal health target. The Lancet Global Health. 2018; 6: e1288–96.

10. Martinez R, Lloyd-Sherlock P, Soliz P, et al. Trends in premature avertable mortality from non-communicable diseases for 195 countries and territories, 1990–2017: a population-based study. The Lancet Global Health. 2020; 8: e511–23.

11. Zhou T, Guan H, Yao J, et al. The quality of life in Chinese population with chronic non-communicable diseases according to EQ-5D-3L: a systematic review. QUAL LIFE RES. 2018; 27: 2799–814.

12. Van Wilder L, Clays E, Devleesschauwer B, et al. Health-related quality of life in patients with non-communicable disease: study protocol of a cross-sectional survey. BMJ OPEN. 2020; 10: e37131.

13. Vos T, Lim SS, Abbafati C, et al. Global burden of 369 diseases and injuries in 204 countries and territories, 1990–2019: a systematic analysis for the Global Burden of Disease Study 2019. The Lancet. 2020; 396: 1204–22.

14. Wang H, Abbas KM, Abbasifard M, et al. Global age-sex-specific fertility, mortality, healthy life expectancy (HALE), and population estimates in 204 countries and territories, 1950–2019: a comprehensive demographic analysis for the Global Burden of Disease Study 2019. The Lancet. 2020; 396: 1160–203.

15. Murray CJL, Aravkin AY, Zheng P, et al. Global burden of 87 risk factors in 204 countries and territories, 1990–2019: a systematic analysis for the Global Burden of Disease Study 2019. The Lancet. 2020; 396: 1223–49.

16. University of Washington, Institute for Health Metrics and Evaluation. Global Burden of Disease (GBD 2019). http://www.healthdata.org/gbd/2019. accessed December 1, 2021

17. Ševcíková H, Raftery AE. bayesPop: Probabilistic Population Projections. J STAT SOFTW. 2016; 75.

18. Raftery AE, Alkema L, Gerland P. Bayesian Population Projections for the United Nations. STAT SCI. 2014; 29: 58–68.

19. United Nations, Department of Economic and Social Affairs, Population Division. Probabilistic Population Projections Rev. 1 based on the World Population Prospects 2019. https://population.un.org/wpp/Download/Probabilistic/Population/. accessed December 1, 2021

20. World Bank. World Bank Country and Lending Groups. https://datahelpdesk.worldbank.org/knowledgebase/articles/906519-world-bank-country-and-lending-groups. accessed December 1, 2021

21. Stevens GA, Alkema L, Black RE, et al. Guidelines for Accurate and Transparent Health Estimates Reporting: the GATHER statement. The Lancet. 2016; 388: e19–23.

22. Riebler A. BAPC: Projection of cancer incidence and mortality data using Bayesian APC models fitted with INLA. https://rdrr.io/rforge/BAPC/. accessed December 1, 2021

23. Riebler A, Held L. Projecting the future burden of cancer: Bayesian age-period-cohort analysis with integrated nested Laplace approximations. BIOMETRICAL J. 2017; 59: 531–49.

24. Schmid VJ, Held L. Bayesian Age-Period-Cohort Modeling and Prediction -BAMP. J STAT SOFTW. 2007; 21.

25. Tsai SP, Lee ES, Hardy RJ. The effect of a reduction in leading causes of death: potential gains in life expectancy. American journal of public health (1971). 1978; 68: 966–71.

26. Eayres D, Williams ES. Evaluation of methodologies for small area life expectancy estimation. Journal of epidemiology and community health (1979). 2004; 58: 243–49.

27. Hu X, Sun X, Li Y, et al. Potential gains in health-adjusted life expectancy from reducing four main non-communicable diseases among Chinese elderly. BMC GERIATR. 2019; 19: 16.

28. Wang GD, Lai DJ, Burau KD, et al. Potential gains in life expectancy from reducing heart disease, cancer, Alzheimer’s disease, kidney disease or HIV/AIDS as major causes of death in the USA. PUBLIC HEALTH. 2013; 127: 348–56.

29. Cieza A, Causey K, Kamenov K, et al. Global estimates of the need for rehabilitation based on the Global Burden of Disease study 2019: a systematic analysis for the Global Burden of Disease Study 2019. The Lancet. 2020; 396: 2006–17.

30. Wang H, Naghavi M, Allen C, et al. Global, regional, and national life expectancy, all-cause mortality, and cause-specific mortality for 249 causes of death, 1980–2015: a systematic analysis for the Global Burden of Disease Study 2015. The Lancet. 2016; 388: 1459–544.

31. James SL, Abate D, Abate KH, et al. Global, regional, and national incidence, prevalence, and years lived with disability for 354 diseases and injuries for 195 countries and territories, 1990–2017: a systematic analysis for the Global Burden of Disease Study 2017. The Lancet. 2018; 392: 1789–858.

32. Anton SD, Woods AJ, Ashizawa T, et al. Successful aging: Advancing the science of physical independence in older adults. AGEING RES REV. 2015; 24: 304–27.

33. Crocker T, Forster A, Young J, et al. Physical rehabilitation for older people in long-term care. COCHRANE DB SYST REV. 2013: D4294.

34. Forster A, Lambley R, Young JB. Is physical rehabilitation for older people in long-term care effective? Findings from a systematic review. AGE AGEING. 2010; 39: 169–75.

35. Abreu A, Pesah E, Supervia M, et al. Cardiac rehabilitation availability and delivery in Europe: How does it differ by region and compare with other high-income countries? EUR J PREV CARDIOL. 2019; 26: 1131–46.

36. Thornicroft G, Ahuja S, Barber S, et al. Integrated care for people with long-term mental and physical health conditions in low-income and middle-income countries. The Lancet Psychiatry. 2019; 6: 174–86.

37. Hearn J, Ssinabulya I, Schwartz JI, et al. Self-management of non-communicable diseases in low-and middle-income countries: A scoping review. PLOS ONE. 2019; 14: e219141.

38. Yusuf S, Joseph P, Rangarajan S, et al. Modifiable risk factors, cardiovascular disease, and mortality in 155 722 individuals from 21 high-income, middle-income, and low-income countries (PURE): a prospective cohort study. The Lancet. 2020; 395: 795–808.

39. Islami F, Goding Sauer A, Miller KD, et al. Proportion and number of cancer cases and deaths attributable to potentially modifiable risk factors in the United States. CA: A Cancer Journal for Clinicians. 2018; 68: 31–54.

40. Grant BF, Chou SP, Saha TD, et al. Prevalence of 12-Month Alcohol Use, High-Risk Drinking, andDSM-IV Alcohol Use Disorder in the United States, 2001-2002 to 2012-2013. JAMA PSYCHIAT. 2017; 74: 911.

41. Cena H, Calder PC. Defining a Healthy Diet: Evidence for the Role of Contemporary Dietary Patterns in Health and Disease. NUTRIENTS. 2020; 12: 334.

42. Afshin A, Sur PJ, Fay KA, et al. Health effects of dietary risks in 195 countries, 1990– 2017: a systematic analysis for the Global Burden of Disease Study 2017. The Lancet. 2019; 393: 1958–72.

43. Licher S, Heshmatollah A, van der Willik KD, et al. Lifetime risk and multimorbidity of non-communicable diseases and disease-free life expectancy in the general population: A population-based cohort study. PLOS MED. 2019; 16: e1002741.

44. Balaj M, Huijts T, McNamara CL, et al. Non-communicable diseases and the social determinants of health in the Nordic countries: Findings from the European Social Survey (2014) special module on the social determinants of health. SCAND J PUBLIC HEALT. 2017; 45: 90–102.

45. Thomson KH, Renneberg A, McNamara CL, et al. Regional inequalities in self-reported conditions and non-communicable diseases in European countries: Findings from the European Social Survey (2014) special module on the social determinants of health. EUR J PUBLIC HEALTH. 2017; 27: 14–21.

46. Collins D, Laatikainen T, Farrington J. Implementing essential interventions for cardiovascular disease risk management in primary healthcare: lessons from Eastern Europe and Central Asia. BMJ Global Health. 2020; 5: e2111.

47. Ganju A, Goulart AC, Ray A, et al. Systemic Solutions for Addressing Non-Communicable Diseases in Low-and Middle-Income Countries. Journal of multidisciplinary healthcare. 2020; Volume 13: 693–707.

48. Tesema AG, Ajisegiri WS, Abimbola S, et al. How well are non-communicable disease services being integrated into primary health care in Africa: A review of progress against World Health Organization’s African regional targets. PLOS ONE. 2020; 15: e240984.

49. Allen LN, Pullar J, Wickramasinghe KK, et al. Evaluation of research on interventions aligned to WHO ‘Best Buys’ for NCDs in low-income and lower-middle-income countries: a systematic review from 1990 to 2015. BMJ Global Health. 2018; 3: e535.

50. Stuckler D. Population Causes and Consequences of Leading Chronic Diseases: A Comparative Analysis of Prevailing Explanations. The Milbank Quarterly. 2008; 86: 273–326.

51. Fidler MM, Gupta S, Soerjomataram I, et al. Cancer incidence and mortality among young adults aged 20–39 years worldwide in 2012: a population-based study. The Lancet Oncology. 2017; 18: 1579–89.

52. Foreman KJ, Marquez N, Dolgert A, et al. Forecasting life expectancy, years of life lost, and all-cause and cause-specific mortality for 250 causes of death: reference and alternative scenarios for 2016–40 for 195 countries and territories. The Lancet. 2018; 392: 2052–90.

